# Exploring Integrated Environmental Viral Surveillance of Indoor Environments: A comparison of surface and bioaerosol environmental sampling in hospital rooms with COVID-19 patients

**DOI:** 10.1101/2021.03.26.21254416

**Authors:** Leslie Dietz, David A. Constant, Mark Fretz, Patrick F. Horve, Andreas Olsen-Martinez, Jason Stenson, Andrew Wilkes, Robert G. Martindale, William B. Messer, Kevin G. Van Den Wymelenberg

**Affiliations:** Biology and the Built Environment Center, University of Oregon, Eugene, OR, 97403; Department of Molecular Microbiology and Immunology, Oregon Health & Science University, Portland, Oregon, USA; Energy Studies in Buildings Laboratory, University of Oregon, Eugene, OR, 97403; Institute for Health and the Built Environment, University of Oregon, Portland, OR, 97209; Healthcare Facilities, Oregon Health & Science University, Portland, Oregon, USA; Division of Gastrointestinal and General Surgery, School of Medicine, Oregon Health & Science University, Portland, Oregon, USA

**Keywords:** SARS-CoV-2, COVID-19, Bioaerosols, Environmental Surveillance, Air Sampling

## Abstract

The outbreak of Severe Acute Respiratory Syndrome Coronavirus 2 (SARS-CoV-2) has dramatically transformed policies and practices surrounding public health. One such shift is the expanded emphasis on environmental surveillance for pathogens. Environmental surveillance methods have primarily relied upon wastewater and indoor surface testing, and despite substantial evidence that SARS-CoV-2 commonly travels through space in aerosols, there has been limited indoor air surveillance. This study investigated the effectiveness of integrated surveillance including an active air sampler, surface swabs and passive settling plates to detect SARS-CoV-2 in hospital rooms with COVID-19 patients and compared detection efficacy among sampling methods. The AerosolSense active air sampler was found to detect SARS-CoV-2 in 53.8% of all samples collected compared to 12.1% detection by passive air sampling and 14.8% detection by surface swabs. Approximately 69% of sampled rooms (22/32) returned a positive environmental sample of any type. Among positive rooms, ~32% had only active air samples that returned positive, while ~27% and ~9% had only one or more surface swabs or passive settling plates that returned a positive respectively, and ~32% had more than one sample type that returned a positive result. This study demonstrates the potential for the AerosolSense to detect SARS-CoV-2 RNA in real-world healthcare environments and suggests that integrated sampling that includes active air sampling is an important addition to environmental pathogen surveillance in support of public health.

## Introduction

A global pandemic was declared 12 March 2020 and is ongoing^1^. Severe acute respiratory syndrome coronavirus 2 (SARS-CoV-2) causes a respiratory illness known as Coronavirus Disease 19 (COVID-19), which can present with a wide variety of symptoms. If symptomatic, the symptoms can mimic the common cold and be extremely mild, or be quite severe requiring medical attention and even hospitalization. In addition to the symptomatic individuals with COVID-19, it is estimated that more than half of all SARS-CoV-2 transmission is due to asymptomatic individuals^2^.

Early in the pandemic, public health officials declared droplet spread as the main route of disease transmission^3^. However, evidence has increasingly implicated inhalation of aerosols in the spread of SARS-CoV-2^4–7^. Past epidemics of coronaviruses, severe acute Respiratory Syndrome-associated Coronavirus (SARS-CoV) and Middle Eastern Respiratory Syndrome-associated Coronavirus (MERS-CoV), demonstrated the ability of virions to spread in built environments via aerosols^8–12^. Previous work has also demonstrated that SARS-CoV-2 can be transported via aerosols and fomites and remain viable in the air and on surfaces^13–15^. Indoor environments with characteristics such as high occupant density, poor ventilation, or high aerosol generating activity have increased potential for transmission of COVID-19, including from pre-symptomatic or asymptomatic individuals^16–19^. Similarly, long-term care facilities (LTCF), hospital care systems, and several other settings that provide services to susceptible occupants are also vulnerable to disease transmission^16,20–24^. The ability to monitor indoor environments to detect potential shedding from infectious individuals is an important layer of control to prevent or contain outbreaks of highly infectious and deadly illnesses like COVID-19.

As a result of the COVID-19 pandemic, there has been increased pressure to innovate current building operational practices, including enhanced air management, and increased motivation to elevate awareness of indoor environmental microbial presence and abundance in general, and specifically to implement ongoing viral surveillance. Environmental surveillance for biological agents is not a new practice. For example, specialized facilities such as military bases and mail distribution centers are monitored for biohazards and biological threat agents such as anthrax and smallpox^25,26^ and water treatment plants monitor for select microorganisms as indicators for drinking water quality^27–30^. Additionally, built environments such as restaurants, food production facilities or hospitals are routinely inspected and sampled for microorganisms to ensure sanitation and cleanliness practices^31,32^. Environmental microbial samples from indoor surfaces are commonly obtained using swabs^33,34^. Aerosol particles can be detected through passive air sampling in settling plates (collection onto the surface of a petri dish)^35,36^ or through active air sampling using a vacuum pump to move a known volume of air across a capture mechanism^37^. Active air samplers span a range of airflow rates, wet and dry media, and physical collection mechanisms including filters, cyclones, impingers and impactors^38–40^. Using any of these collection devices, the sampling media can be evaluated for a target pathogen using molecular techniques.

SARS-CoV-2 environmental sampling is currently being implemented on college campuses (wastewater) and in LTCFs (surface swabs) to identify and prevent potential outbreaks in high density living situations^20,41–44^. Wastewater surveillance is suited to detect the presence of SARS-CoV-2 in a large geographic region, possibly down to the scale of a cluster of buildings or a single building, but less likely to provide spatial resolution down to specific zones or rooms within a building^45,46^. Furthermore, not all occupants use restrooms while occupying public buildings, increasing the possibility of missing infected occupants. Surface swabs are suited to capture a time integrated exposure within a space, but effectiveness may vary depending on cleaning practices, and may not detect viral RNA that remained suspended in aerosols long enough for the room air to be exchanged from a space before settling could occur. Moreover, based on room air fluid dynamics, uneven spatial settling likely occurs^47^ and depending on the number and spatial resolution of surface swab or settling plate samples collected, these methods may not reflect the dynamic aerosol viral load. Each method of sampling has strengths and limitations, and environmental surveillance may ultimately be most effective if implemented in an integrated fashion; however, at present, little surveillance has been routinely conducted via bioaerosol sampling. With the growing evidence that COVID-19 is spread through virus-containing aerosol emissions and the known limitations of other environmental surveillance techniques of SARS-CoV-2, it may be beneficial to supplement these with a sensitive and robust bioaerosol sampling platform. To explore this question, we initiated a field study within a healthcare environment including environmental sampling via surface swabs, settling plates, and an active air sampler to explore the relationships of these sampling approaches.

As an environmental sampling testbed, healthcare facilities provide an excellent opportunity to identify surface and aerosol contamination in rooms where viral particles may be emitted from patients^11,48–55^, while also a suitable challenge due to enhanced decontamination protocols. Among built environments, healthcare facilities have some of the most advanced strategies to reduce pathogen transmission risk indoors, including high air exchange rates^56^, high filtration efficiency, isolation environments and access to personal protective equipment (PPE), and established hand hygiene practices^57^. Nonetheless, current literature suggests that transmission stemming from hospital-associated infections range from 14.1%-41% of in-house COVID-19 cases^58–60^, further justifying healthcare facilities as an environmental surveillance testbed.

Horve et al. conducted benchtop and room-scale experiments to determine the feasibility of air sampling as an environmental surveillance tool^61^. Specifically, they reported that AerosolSense (Thermo Fisher Scientific, Catalog #2900-AA) had robust detection capability for heat-inactivated SARS-CoV-2 at aerosol viral concentrations of ~30 genome copies per liter (gc/L) of room air for 75 minute sampling periods and as little as ~0.01 gc/L of room air for >8 hour sampling periods^61^. From November to December 2020, we collected environmental surface swabs and passive air settling plates from COVID-19 patient rooms at Oregon Health & Science University (OHSU) Hospital in Portland, Oregon. Simultaneously, we collected air samples using the AerosolSense. The objective of this study was to determine effectiveness of the AerosolSense air sampler to detect SARS-CoV-2 and to better understand the relationship between air and surface sampling in the built environment.

## Methods

Environmental samples were collected from COVID-19 patient rooms (n=32) at OHSU from November 2020 to December 2020. Factors for choosing patient rooms were severity of illness, type of oxygen support and planned or anticipated aerosol generating procedures. These COVID-19 positive patients were determined through either an initial rapid SARS-CoV-2 antigen test followed by a RT-PCR diagnostic test or a RT-PCR diagnostic test only. COVID-19 positive patients were housed in wards 5A (Acute Care), 5C (Family Medicine), 8D (Emergency Department), 7A (MICU), 12C (Labor and Delivery), and 14C (Internal Medicine Inpatient). All OHSU PPE donning, doffing and safety procedures were strictly followed to prevent contamination and illness to healthcare workers (HCW) and/or researchers.

### Bioaerosol sampling

AerosolSense instruments were calibrated to sample 200 liters per minute (L/min) of room air across AerosolSense Capture Media (ACM) using the Streamline Pro MultiCal System (Chinook Engineering, Wyoming). The air sampling devices were deployed in COVID-19 patient rooms and allowed to run for at least one hour, with the majority of sampling events lasting in excess of two hours. After sampling, the ACM was placed into a lysis/preservative buffer (DNA/RNA Shield, Zymo Research #R1100) for immediate preservation of nucleic acids, and the sampling run time was recorded. After each sampling event, the device was decontaminated using Cavicide (Metrex), allowing a 2 minute period for the Cavicide to inactivate microorganisms, and then removed from the patient room. The device was then cleaned again using Sani-Cloth Bleach Germicidal Wipes (PDI #U26595).

### Surface Swab and Settling Plate Sampling

At the end of the air sampling duration, surface swab samples were taken from multiple surfaces in COVID-19 patient rooms using 15mL conical tubes (Cole-Parmer UX-06336-89) and polyester flocked swabs (Puritan #25-3060-H) pre-moistened with Viral Transport Media (VTM) (Rocky Mountain Biologicals) from the tube. A predesignated sampling area was swabbed in an overlapping “S” pattern, first horizontally then vertically, to ensure complete coverage of the area. The moistened swab was also rotated during collection so that optimal surface area of the swab was used for sample collection and then returned to the conical tube with the remaining VTM. The sampling area was approximately 20 cm by 30 cm on patient room surfaces and included: work counter, return air grille, supply air grille, hopper sink, floor patient left, floor patient right, floor patient foot, floor patient head (when possible), lavatory floor, lavatory exhaust air grille and hallway floor immediately outside patient room. In ICU rooms, there is no designated lavatory, and the sluice sink was swabbed instead. After taking a sample, the swab was then returned to the 15mL conical tube containing the remainder of VTM. Sample tubes were placed on ice in a designated sample cooler until processing. Settling plate samples were collected from work counters, windowsills, supply carts, nurses stations, under the patient’s bed, and various locations on the floor of the occupied rooms. Standard petri dishes (100 mm x 15 mm) were set out with both halves open for the entire active air sampling duration. At sample collection, the inside surface of both halves of the petri dish were swabbed as described above.

### Sample processing

The sample cooler was hand carried to a BSL-2+ lab on the OHSU campus for processing. All sample processing was conducted in a class-2 biosafety cabinet (BSC). The environmental surface samples (flocked swabs and passive settling plates) previously placed in 15 ml conical tubes were vortexed briefly and then incubated at room temperature for five minutes. A 200 µL aliquot of the supernatant was removed and placed into a microcentrifuge tube (Thomas Scientific #1223K29) containing 600 µL of lysis/preservative buffer (DNA/RNA Shield, Zymo Research #2100). Samples were then transported by automobile to a BSL-2 laboratory on the University of Oregon campus in Eugene, Oregon, USA. Total RNA was extracted from all samples using Zymo Quick-DNA/RNA Viral MagBead kit (Zymo Research #R2141) and stored at −80°C until analysis. Successful RNA extraction was confirmed using a 5 uL spike-in of *Escherichia coli MS2* bacteriophage that was added to each reaction mixture.

### Molecular Analysis

SARS-CoV-2 RNA presence and abundance was determined by qRT-PCR. The TaqPath COVID-19 Combo Kit (Thermo Fisher Scientific, Catalog #A47814) targeting the N, S, and ORF1ab (RdRP) gene regions was used to prepare qRT-PCR reactions for processing. Each reaction mixture contained 5 µL TaqPath 1-Step Multiplex Mastermix without ROX (Thermo Fisher Scientific, Catalog #A28521), 9 µL nuclease-free water (Invitrogen, Catalog #4387936), 1 µL COVID-19 Real Time PCR Assay Multiplex Mix (Thermo Fisher Scientific, Catalog #A47814), and 5 µL of extracted RNA. Thermocycling was performed with the QuantStudio5 (Applied Biosystems) using the following cycling conditions: 25°C for 2 minutes, 53°C for 10 minutes, 95°C for 2 minutes, and 40 cycles of 95°C for 3 seconds and 60°C for 30 seconds. If the presence of SARS-CoV-2 RNA was detected, and the cycle threshold (C_t_) was less than 35, with observed amplification in two out of the three genome targets, then a sample was considered positive. This follows the FDA Emergency-Use Authorization guidelines in the assay instructions for use^62^. Two control samples were included in the qRT-PCR reaction. These consisted of an extraction control from the RNA extraction process and a non-template control (NTC) to account for possible laboratory contamination. Reagent controls were processed concurrently with environmental samples. All controls tested negative for the presence of SARS-CoV-2 RNA.

### Institutional Approval and Data Availability

The research described was determined to be IRB exempt and granted an IRB exemption. This work was reviewed by the OHSU Institutional Biosafety Committee and approved under PROTO202000016. Data and analysis scripts are available on GitHub (https://github.com/BioBE/AerosolSense-FieldTrials).

### Statistical Analyses

All analyses were performed using the statistical computing environment, R^63^. Differences between samples from the same room were computed using paired t-tests. Differences were considered significant with *P < 0*.*05*.

## Results

The overall objective of this investigation was to explore integrated environmental surveillance and to determine the potential efficacy of the AerosolSense active air sampler for the detection of SARS-CoV-2-containing aerosols in a real-world setting. To this end, 39 aerosol samples collected, 132 passive air samples were collected in settling plates, and 317 surface samples were collected with flocked swabs from 32 COVID-19 patient rooms at OHSU over a two month sampling period. All sampling locations were assessed for the percent of samples that returned a positive result for the presence of SARS-CoV-2 (Figure 1). Positive samples were returned for 53.8% of air samples,12.1% of passive settling plates, and 14.8% of room surface swabs. After the active air samples (53.8% positive), the most frequent positive sample locations were swabs taken from the floor to left side of the patient bed (25.0%), the hallway floor directly outside the patient room (~21.4%), settling plates in the hallway outside the patient rooms (20.0%), and the lavatory exhaust air grille (20.0%).

**Figure 1.**
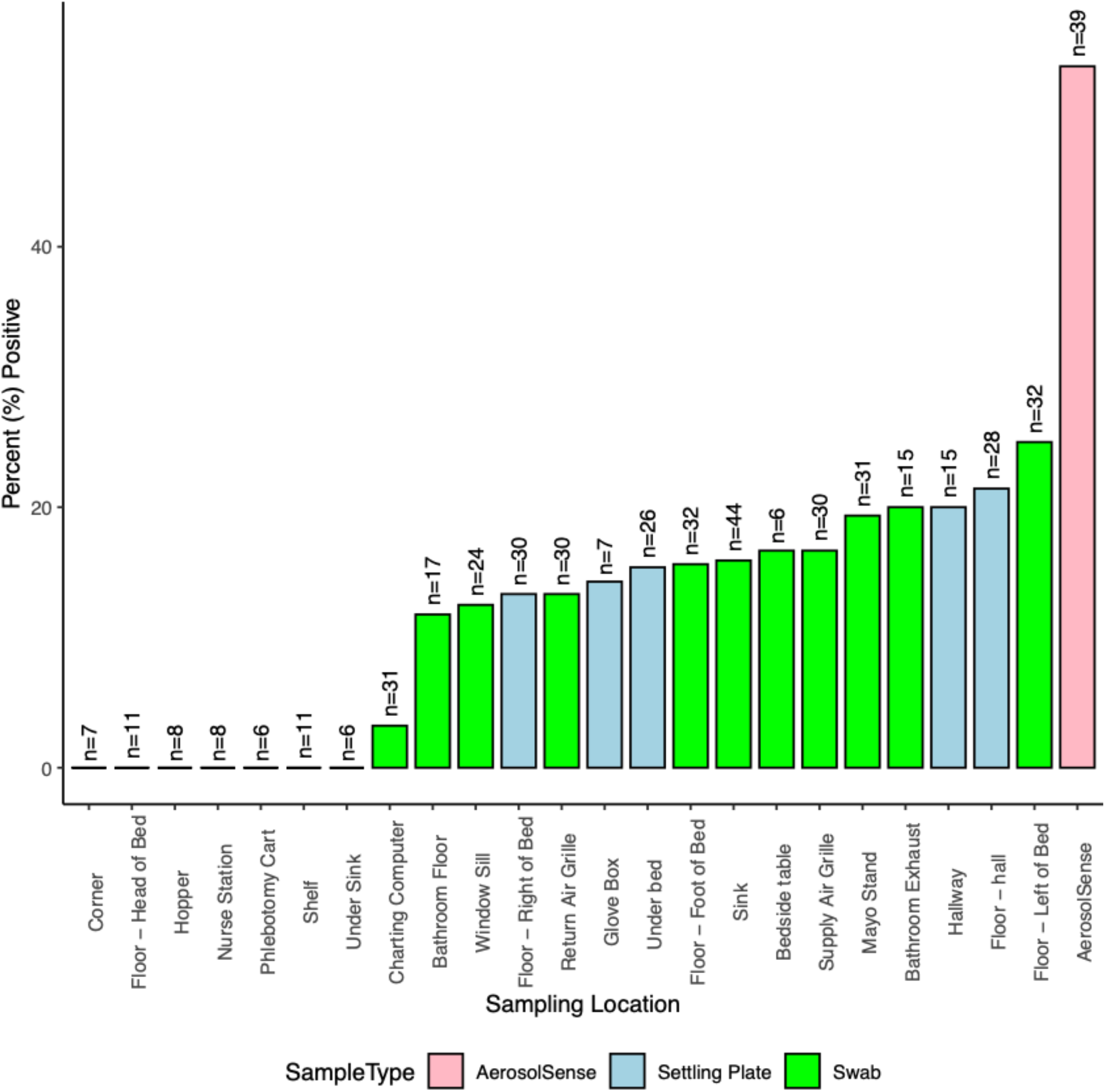
Percent of samples found to be positive for the presence of SARS-CoV-2 RNA at each location sampled. The number of total samples collected are listed at the top of each bar. A full description of each sampling location can be found in the supplementary data. Pink indicates the active air sampler, blue indicates the passive air settling plates, and green indicates surface swabs. Sampling locations with fewer than seven (n=7) were excluded from this figure.

Overall, all three sampling types (active air, swabs, settling plates) were able to successfully isolate SARS-CoV-2 RNA (Figure 2). In order to assess the potential for genomic material capture differences, and ultimately to better understand sampling methods for environmental viral surveillance, sampling method was compared for all rooms that returned any positive result whatsoever. In total, about 69% (22/32) of sampled rooms had a positive environmental sample of any type. Among these rooms, 32% (7/22) had only the air samples that returned positive, 27% (6/22) had only one or more surface swabs that returned positive, 9% (2/22) had only passive settling plates return a positive result, and 32% (7/32) had multiple sample types that returned positive. Additionally, among the 7 rooms that returned positive for both air and surface swab(s), positive samples collected by the air sampler were found to have significantly lower C_t_ values (Paired t-test; *P < 0*.*05*) than positive environmental surface swabs (Figure 3).

**Figure 2.**
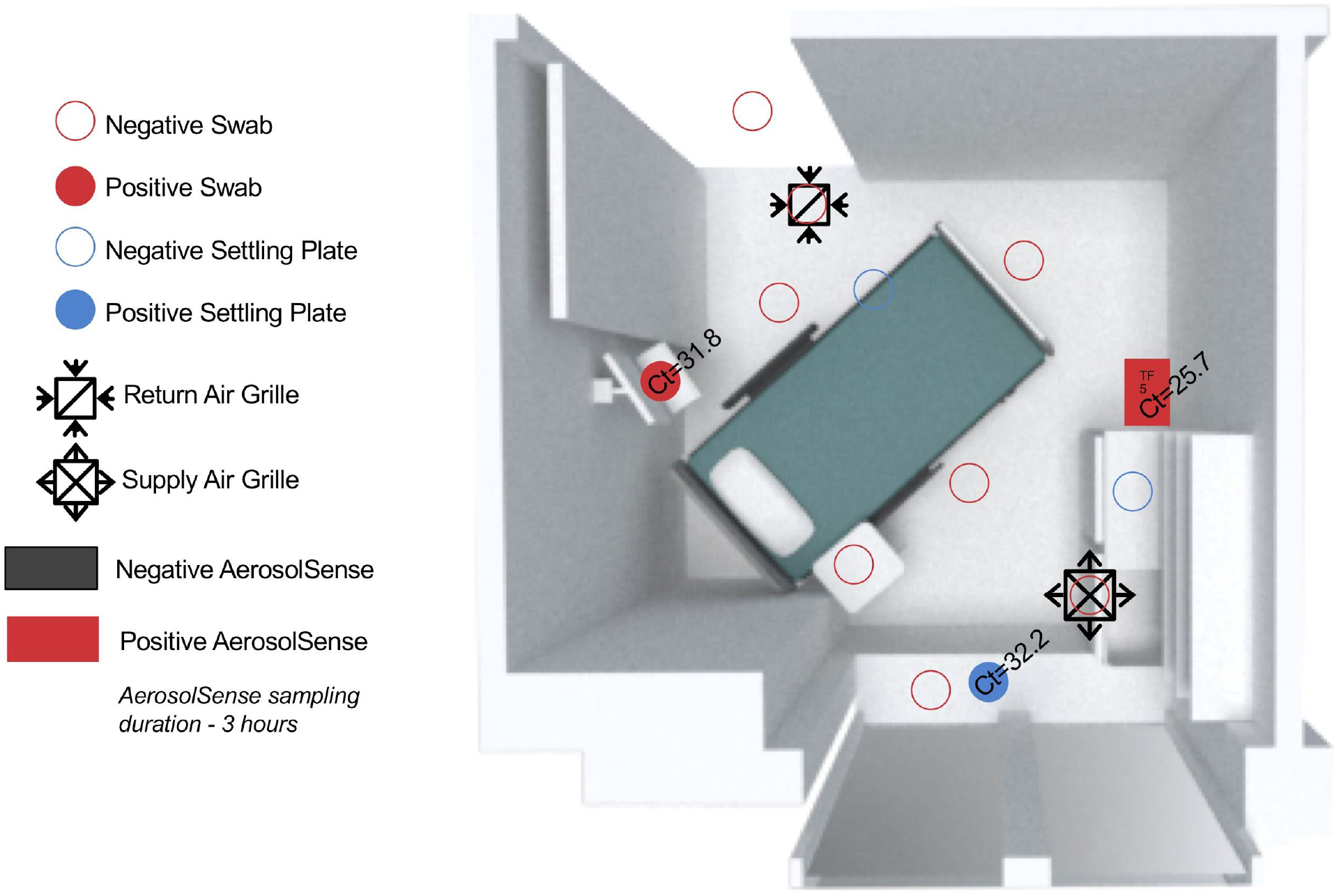
Room plan of one sampled patient room indicating sampling type, number and location. Cycle threshold (Ct) values from qRT-PCR test results are included, with lower values indicating higher abundances of SARS-CoV-2 RNA. Rectangles indicate AerosolSense samplers, blue circles indicate passive air settling plates, and red circles indicate surface swab locations. Filled circles indicate a positive result and open circles indicate a negative result. This room had a positive active air sample, charting computer swab, and windowsill settling plate.

**Figure 3.**
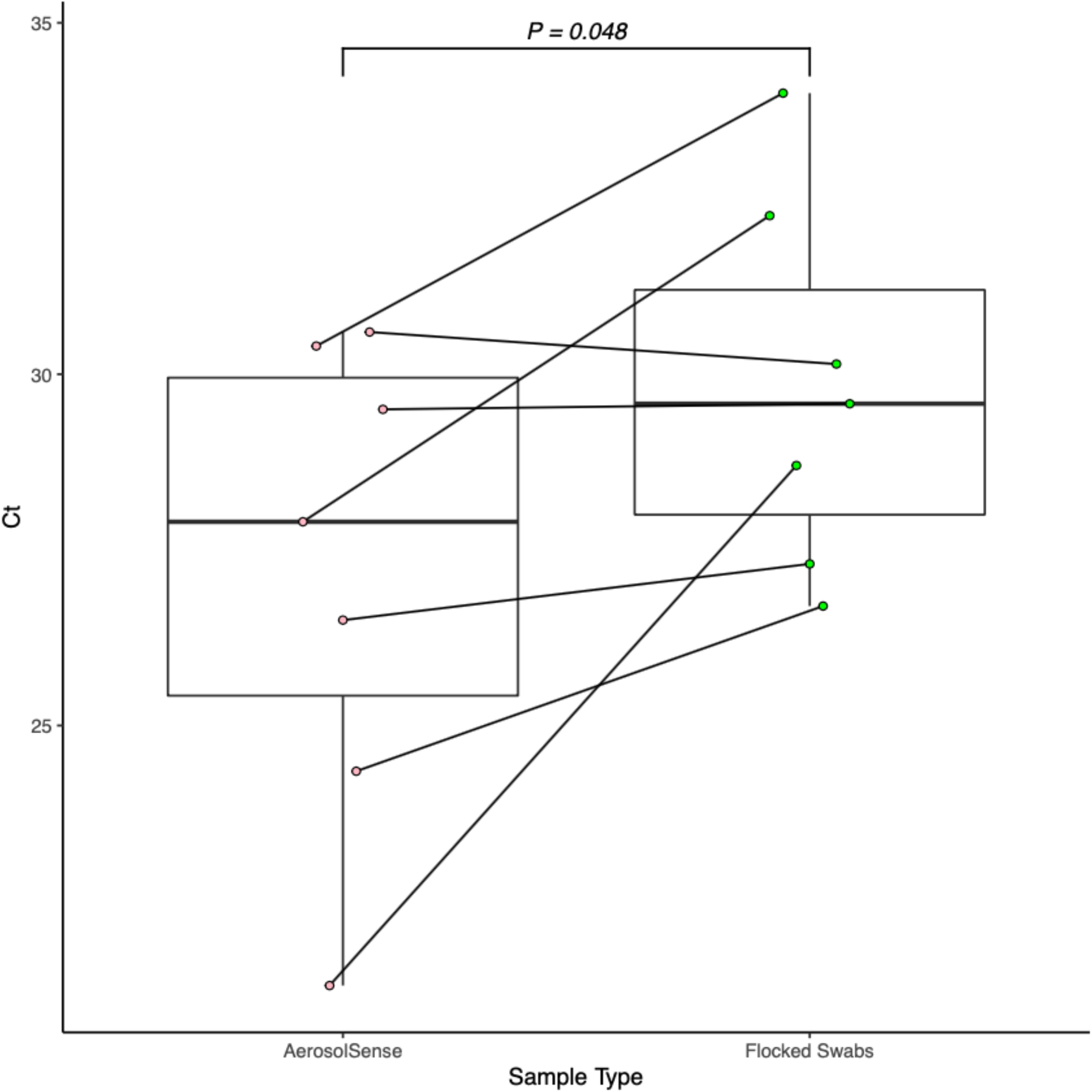
Box and whisker plot demonstrating the minimum, maximum, median, and quartiles C_t_ values observed in samples recovered from the AerosolSense sampler or environmental swabs, among rooms that returned positive for both sampling types. Lines connect the mean Ct value of positive samples taken from the same patient room at the same sampling time. Positively sloped lines indicate a lower C_t_ value (higher abundance) of SARS-CoV-2 RNA in air samples. Pink points are the mean (if applicable) of the Ct values of samples collected with the air sampler and green points are the mean of all the positive surface swabs from a given room.

## Discussion

This study contains a few important limitations. Aerosol sampling durations were variable (1-12.75 hours) in order to prioritize the schedule and activities of the care team providing patient care. Approximately 81% of study rooms (26/32) had one air sampler while approximately 19% of the rooms (6/32) had two air samplers during the sampling period. All rooms with two air samplers returned the same result for both samplers. Verified hospital room air exchange rate was not available for every patient room studied. The analysis methods cannot ensure that RNA detected in any room came solely from the patient occupying the room. Finally, our results report the presence of SARS-CoV-2 RNA, and do not address viability, since qRT-PCR does not distinguish viable virions from RNA from non-viable cells.

Overall, we sought to identify the potential utility of the AerosolSense device for active air environmental surveillance and used a healthcare setting as the testbed. The active air samples recovered were compared to the current standard for indoor environmental surveillance, flocked environmental swabs and passive air settling plates^64–67^. We confirmed the results of previous studies that demonstrated significant environmental contamination by SARS-CoV-2 in rooms occupied by COVID-19 positive patients^14,50,54,64,68^. While one may expect more consistent detection of SARS-CoV-2 RNA in rooms occupied by COVID-19 positive patients, shedding has been shown to vary by individual and decrease, even as symptoms and disease progresses^69^ and our results are consistent with other investigations^14,54^. Furthermore, at this facility patient rooms maintain at least six air changes per hour (sometimes much higher), COVID-19 patients are placed into negative pressure isolation rooms whenever possible, and all COVID-19 patient rooms undergo rigorous daily cleaning protocols, all with the intention of reducing overall environmental contamination.

Our data demonstrated significantly lower C_t_ values in samples collected from the air sampler compared to environmental swabs, among rooms where both sampling methods returned a positive result. The lower paired C_t_ values observed in the air sampler and the higher percentage of positive air samples (~54%) when compared to other sampling methods, along with its ability to detect SARS-CoV-2 in some rooms where other methods did not (32%), suggests that bioaerosol surveillance of SARS-CoV-2 makes an important contribution to environmental viral surveillance techniques. Nonetheless, there was reasonable concordance between active air samples and surface swabs, with both sampling methods signaling room contamination for 32% of rooms that tested positive. As stated, there was a meaningful additional percentage (32%) of positive rooms were only detected via active air sampling, while 27% of positive rooms were only detected via surface swabs, thus supporting the value of integrated surveillance. Surface swabs and settling plate collection methods benefitted from greater number and spatial resolution and sample number, while the active air sampler benefitted from continuous sampling and spatial integration via mixing of room air. Furthermore, surface swab samples capture a time-integrated history of direct contact and particle deposition that occurred since previous decontamination, while active air samples represent a specific sampling duration, volume of air, and have an opportunity to capture particles that do not deposit onto surfaces.

Overall, the air samples had the most prevalence (by percentage) of detecting SARS-CoV-2. The patient room sampling locations that had the second most prevalence of detecting SARS-CoV-2 were surface swabs that were collected near the patient (floor adjacent to bed and mayo stand) and from areas where SARS-CoV-2 was likely sourced through aerosols (patient room return air grille and lavatory exhaust air grille). It is expected that areas nearer the patient (such as the mayo stand, bedside table, and floor samples) would exhibit surface contamination, however the prevalence of return and exhaust air grilles is less commonly reported^54,70^. The contamination observed in hallways (swabs 21.4% and settling plates 20.0%) was possibly sourced from within the adjacent patient room and further spread by airflow, HCW foot traffic, or the movement of equipment carts necessary for care^71,72^, or may have been sourced from outside the patient room. We observed viral contamination on low-touch surfaces (return air grilles, supply air grilles, and windowsills). These locations are beyond the expected range of routine droplet transport, rarely come into contact with individuals, and may not be routinely decontaminated. The supply air grilles that tested positive (16.7%) may have been contaminated with SARS-CoV-2 from recirculation of building ventilation air^73^, from non-laminar flow of supply air out of the grille, or potentially form within-room surface deposition or impacation sourced from high velocity droplet generating events.

The presence of SARS-CoV-2 RNA in the active air samples and upon surfaces that are commingled with active room airflow (supply, return, and exhaust air grilles), combined with the growing evidence of the potential for aerosol-based disease transmission^16,20,53,66,73–81^, presents a compelling argument for the merit of indoor air microbial surveillance. Moreover, due to the spatially integrated nature of indoor aerosols, continuous air sampling techniques with sufficient sensitivity can be incredibly useful to increase situational awareness and guide building operational improvements to reduce indoor disease transmission risk^82^.

When encountered with a possible infectious disease outbreak, epidemic or pandemic, healthcare facilities and government public health agencies respond with containment strategies. This response strategy is well documented by most global governing bodies with a structured healthcare system. The steps of infectious agent containment include: 1) identification of agent 2) infection control assessment 3) health screenings where appropriate 4) coordinated response efforts and 5) continued assessments and health screenings until containment is achieved^83^. Built environment surveillance in general, and active air monitoring in specific, should be an integral and proactive component of a comprehensive infectious disease management strategy. By pairing these surveillance data with appropriate building operations layered risk reduction strategies, the transmission of disease indoors can be minimized and potentially avoided.

## Conclusion

Currently, the majority of environmental surveillance for microorganisms utilize wastewater and surface sampling. Wastewater, surface swabs, and aerosol surveillance methods each have strengths and limitations, and are best implemented in an integrated manner. Wastewater sampling provides excellent insight to larger geographic scales disease prevalence but has limitations for guiding actions within a specific facility. Surface samples are time-integrated and are influenced by decontamination protocols, as well as spatial resolution and room air dynamics. This research demonstrates the added detection capability of bioaerosol sampling in environmental viral surveillance. Specifically, this research demonstrates that the AerosolSense active bioaerosol sampling platform effectively detects SARS-CoV-2 RNA in a real-world healthcare environment with high air exchange rates.

## Data Availability

Data and analysis scripts are available on GitHub (https://github.com/BioBE/AerosolSense-FieldTrials).

## Acknowledgements

Gratitude for the Biology and the Built Environment Laboratory students Liliana Barnata and Vincent Moore, who assisted with sample collection on-site. A special thanks to staff Georgia MacCrone and Surbhi Nahata, who assisted with the lab work. The authors would like to thank Siqi Tan, Arunava Dutta, and Geoffrey Gonzalez for their review of the manuscript.

Lastly, special recognition for the many frontline health care workers, who provide tireless and compassionate care for their patients.

